# The association between the number of Antenatal Care (ANC) visits and neonatal mortality in Afghanistan: a cross-sectional study based on the 2015 Demographic and Health Survey

**DOI:** 10.1101/2025.10.06.25337466

**Authors:** Fida Khallouf, Sam Newton

## Abstract

**Background:** Neonatal mortality in Afghanistan is among the highest worldwide, driven by rural–urban disparities, low maternal education, and poor healthcare access. Antenatal care (ANC) remains severely underutilized, with fewer than one in five women meeting minimum visits despite its link to neonatal survival, highlighting the need to assess its impact in conflict-affected settings.

**Objectives:** Determine whether the number of antenatal care visits during pregnancy is associated with neonatal mortality in Afghanistan and investigate whether 4 ANC visits could be a sufficient minimum requirement to prevent neonatal mortality in conflict settings such as Afghanistan.

**Methods:** Analytic, population-based, cross-sectional study. Using data from the 2015 DHS for Afghanistan, logistic regression was performed to assess the association between fewer than 4 ANC visits and neonatal mortality.

**Results:** The overall prevalence of neonatal mortality was 1.85% (CI:1.56-2.20). The odds of neonatal mortality for children whose mothers attended ANC less than 4 times were 1.8 times higher in comparison to those whose mothers attended ANC 4 times or more. After adjusting for the age of the mother, sex, ANC provider, place of residence (urban/rural), wealth index, education of mother, and education of father, the odds of neonatal mortality were 1.1 (95% CI 0.72-1.71) with not enough evidence against the null hypothesis (p-value 0.637). While the mother’s education, place of residence, wealth index, ANC provider, and father’s education confounded the relationship, sex showed an interactive effect on the main relationship.

**Conclusions:** ANC visits less than 4 times were 1.8 times more likely to result in neonatal mortality than ANC visits 4 or more times. This study confirmed the importance of setting 4 visits as a minimal threshold for improved pregnancy outcomes, even where access to health care could be challenging, such as in Afghanistan.

## 1. Introduction

This project assesses whether conducting four or more antenatal care visits is associated with decreased neonatal mortality in Afghanistan. The World Health Organization defines neonatal mortality as the deaths among live births during the first 28 completed days of life (1). Neonatal mortality in Afghanistan decreased from 45 deaths per 1000 live births (2006–2010), which was substantially higher than the global average of infant mortality (32 per 1,000 live births) (2), to 35.5 deaths per 1000 live births in 2022 (3).

Despite a 44 % decline in neonatal deaths since 2000, nearly half (47%) of all deaths under 5 years in 2022 were in the neonatal period, which is among the most vulnerable periods of life and requires intensified quality intrapartum and newborn care (4).

Substantial global progress has been made in reducing childhood mortality since 1990. The total number of under-5 deaths worldwide has declined from 12.8 million in 1990 to 4.9 million in 2022, and the global under-5 mortality rate has dropped by 59%, from 93 deaths per 1000 live births in 1990 to 37 in 2022. The global number of neonatal deaths has also declined, from 5.2 million in 1990 to 2.3 million in 2022. However, the decline in neonatal mortality from 1990 to 2022 was slower than that of post-neonatal under-5 mortality, according to the UNICEF data (5). There are approximately 6,300 newborn deaths every day, amounting to nearly 47% of all child deaths under the age of 5 years (6).

In the 5 years preceding the 2015 Afghanistan Demographic and Health Survey (AfDHS), the neonatal mortality rate was 22 deaths per 1,000 births, meaning that one of every 45 children died during the first month of life. Two-fifths of deaths below the age of 5 occurred during the first month of life (7). While these mortality estimates appear lower than expected, conflict-related challenges in data collection suggest underreporting, and therefore, these estimates should be cautiously interpreted. According to the AfDHS, factors such as size at birth and sex influence neonatal mortality. The neonatal mortality rate is 35 deaths per 1,000 live births among neonates reported to be smaller than average at birth, as compared with 16 deaths per 1,000 live births among those who were of average or larger size at birth. Furthermore, boys were more likely to die within the first 28 days than girls. Neonatal mortality rates had shown a decline over the years preceding the DHS; the estimates declined from 31 to 28 to 21 per 1000 births over the periods 14-10, 5-9, and 0-4 years, respectively, before the survey. However, this decline was associated with conflict-related impacts on data collection and reporting, as neonatal death, in particular, appears to be under-reported. Further analyses are being conducted to understand these estimates better, and additional analyses are encouraged. Considering that the perinatal mortality rate encompasses both stillbirths and early neonatal deaths, determinants affecting the perinatal rate could subsequently have an impact on neonatal death.

Maternal and reproductive health is one of the core components to achieve the 2030 agenda for Sustainable Development Goals (SDGs), particularly target 3, which seeks to ensure healthy lives, promote well-being for all ages, and reduce neonatal mortality to less than 12 per 1000 live births (8). Adherence to ANC has proven to contribute to reduced childhood mortality and morbidity. A study investigating the association of ANC in low-income and middle-income countries with short- and long-term mortality and nutritional child outcomes concluded that at least one ANC visit was associated with 1.04% points reduced probability of neonatal mortality and having at least four ANC visits and having at least once seen a skilled provider reduced the probability by an additional 0.56% and 0.42% points, respectively (9).

It is widely accepted that a good quality antenatal care service during pregnancy will reduce the risk of neonatal mortality, and the World Health Organization has increased the recommended number of ANC visits from four to eight based on recent evidence that a higher frequency of ANC contacts with a health provider is associated with a reduced likelihood of stillbirths (10). In low- and middle-income countries such as Afghanistan, much effort is yet to be made to reduce neonatal mortality. Due to a prolonged period of conflict and war, Afghan women are currently struggling to reach ANC services through which, in some situations, either their safety could be jeopardized or they might risk losing financial assets. The low uptake of at least four ANC sessions underscores a critical issue; there is little to gain in pushing for an increase to eight visits when the existing structure fails to retain women through even half of the recommended visits (11). The analysis in this project is aimed at evaluating whether there is a significant impact on neonatal mortality if a woman attends ANC services less than 4 times and, consequently, whether there is a need in the Afghanistan context to increase adherence to a minimum of 4 ANC visits during pregnancy.

### 1.1 Background

#### 1.1.1 Literature review

Rural versus urban healthcare disparities, prenatal and postnatal care quality, and the influence of healthcare infrastructure have been identified in previous literature and through clinical practice as main determinants of neonatal mortality in which antenatal care visits play an integral role (12). Huge disparities in neonatal mortalities have persisted across regions, with Afghanistan being amongst the countries with the highest rates alongside Somalia, South Sudan, and Pakistan (13).

The AfDHS for 2015 identified the mother’s age at birth, short birth intervals, rural areas (twice as high as urban areas), and household wealth as the most prominent determinants of neonatal mortality. The neonatal mortality rate increased from 17 to 27 per 1000 live births in rural areas compared to urban areas, and from 18 in those whose mothers had at least primary education to 26 in those whose mothers had no education (7).

Although there have been multiple studies investigating the determinants of neonatal mortality in Afghanistan, there has been little focus on addressing the root causes of low ANC adherence in the conflict-affected context of Afghanistan. It was discovered in a study analyzing the 2015 DHS of Afghanistan that more than one-third of the women did not receive a single antenatal care visit during pregnancy, indicating that this important service is severely under-utilized in this country (11). Data from 2015 reveals that only 59% of pregnant women accessed ANC services at least once during their pregnancy. Despite WHO’s recommendation of eight ANC visits, in 2018, only 21% of Afghan pregnant women accessed four ANC sessions (14).

In another study analyzing household surveys from 60 low- and middle-income countries, a new indicator to measure quality ANC was proposed: ANC q or the adequacy score, which was concluded as low (3.5) in Afghanistan in comparison to other countries (15). Considering the protective effects of quality ANC services on neonatal survival and the low adequacy score for ANC in Afghanistan, there is a need to investigate the true effect of the number of ANC visits on neonatal mortality in the context of Afghanistan. Conflict-affected settings could benefit from rationalizing the requirements for the minimal number of ANC visits needed to prevent neonatal mortality while being mindful of the challenges that might hinder pregnant women living in these regions from accessing and adhering to ANC visits.

Furthermore, for maternal health services such as institutional delivery and antenatal care, place of residence is a major factor explaining inequality of utilization among the population. Socioeconomic status, availability of transportation, and accessibility of health-related information all contribute to the inequality of access to health services in Afghanistan (7).

According to AfDHS 2015, fifty-nine per cent of women who gave birth in the 5 years before the survey received antenatal care from a skilled provider for their most recent birth. However, only 18% had the recommended four or more ANC visits. Women in urban areas are more likely to receive ANC than women in rural areas (72% versus 55%). Also, urban women are more likely than rural women to receive ANC from a doctor (46% versus 26%), and ANC coverage differs substantially by province. The AfDHS also showed that there is a strong association between ANC visits and institutional deliveries; specifically, the more ANC visits, the better the chance of an institutional delivery that improves neonatal mortality in high-risk pregnancies. Seventy-eight per cent of births among mothers with four or more ANC visits were delivered in a health facility, as compared with only 30% of births among mothers with no ANC visits, concluding that institutional deliveries were also less common in rural areas, increasing the risk of neonatal mortality in high-risk pregnancies.

Additionally, there was a strong association between mothers’ education and timely newborn postnatal checkups. Only 9% of newborns whose mothers have no education receive a postnatal checkup, as compared with 22% of newborns whose mothers have more than a secondary education, further stressing the importance of mothers’ education as a predisposition for adequate health care for neonates. Regarding accessing healthcare, most of the problems mentioned in the DHS report were in rural areas, further demonstrating the disparities between rural and urban areas.

### 1.2 Aims and Objectives

This study aims to assess the impact of poor adherence to antenatal care visits on pregnancy outcomes such as neonatal mortality.

Objectives:

1. To assess whether neonatal mortality is higher when mothers attended antenatal care services less than 4 times.
2. To understand how many ANC visits women in a conflict setting in Afghanistan are likely to conduct.
3. To identify whether 4 ANC visits could be a standard minimum threshold for preventing neonatal mortality.

### 1.3 Null hypothesis

H0: There is no difference in the likelihood of neonatal mortality between women who attend ANC less than 4 times during pregnancy and those who attend ANC four or more times during pregnancy.

## 2. Materials and methods

The research question was answered by conducting a statistical analysis using logistic regression to assess the association between the exposure of interest (less than 4 ANC visits) and the outcome of interest (neonatal mortality) in comparison to those who conducted 4 or more ANC visits during their pregnancy. Data on neonatal mortality was obtained from the DHS survey referring to the last child born and on which the ANC data is collected. The crude association was assessed as a starter. It was followed by adjusting for possible confounders and effect modifiers such as the age of the mother, sex, marital status, education of the mother, urban/rural residence, smoking status of the mother, where ANC care was attended, ANC care attended by a health practitioner, socioeconomic status (using wealth index in the Afghanistan Mortality Survey 2010), and education of the father.

The exposure of interest was categorized into a binary numerical variable with a cut-off point of 4 ANC visits. The outcome was also categorized into a binary variable (alive or dead within the first 30 days after birth). The crude association between less than 4 ANC visits and neonatal mortality was measured using risk ratios. Further, odds ratios of neonatal mortality will be measured by comparing those who visited less than 4 times and those who visited 4 or more times.

The dataset used is from the general survey for 25,974 enumeration areas of a stratified sample with a ratio of rural to urban representation of 78: 23%. This general survey is sufficient for the research question and has suitable data for the project. The DHS sample excluded areas with ongoing security issues and replaced them with other chosen clusters within the province without exceeding 10% of the selected clusters. Probability proportional to the size of the enumeration area was considered while dividing large areas into segments. Therefore, in 2015, DHS for Afghanistan, which in total has 956 clusters, a cluster is either an enumeration area or a segment of it. Due to the nonproportional allocation of the sample of the DHS across provinces, sample weights was used for the analysis.

### 2.1 Study design

This is an analytical, population-based, cross-sectional study using data collected for the 2015 Demographic Health Surveys of Afghanistan, which was accessed for the purpose of this study on the 24^th^ of November 2023. The purpose of the DHS survey is to collect demographic and health data from different populations at regular intervals using a standardized methodology for comparison between populations over time (16).

### 2.2 Study setting

A total of 198 fieldworkers were required to collect data for the 2015 DHS of Afghanistan. They were part of the group of 300 individuals trained in Kabul by the Central Statistics Organization and the DHS program trainers to conduct the fieldwork. All data collection took place via home visits across the country between June 15, 2015, and February 23, 2016.

### 2.3 Study population

The study included all women aged between 15 and 49 who had spent the previous night in the household and provided birth and pregnancy history.

#### Sampling frame

The sampling frame used for the 2015 AfDHS had information from 25,974 enumeration areas (EAs). It followed a stratified two-stage sample design intended to allow estimates of key indicators at the national level, in urban and rural areas, and for each of the 34 provinces of Afghanistan.

#### Sampling method

- Stage 1: select sample points (clusters) consisting of EAs. A total of 950 clusters were selected, 260 in urban areas and 690 in rural areas. It was recognized that some areas of the country might be difficult to reach because of ongoing security issues. Therefore, to mitigate the situation, reserve clusters were selected in all of the provinces to replace the inaccessible clusters. The 101 reserve clusters that were preselected did not exceed 10% of the selected clusters in the province.
- Stage 2: systematic sampling of households. A household listing operation was undertaken in all of the selected clusters, and a fixed number of 27 households per cluster were selected through an equal probability systematic selection process for a total sample size of 25,650 households. Because of the approximately equal sample size in each province, the sample is not self - weighting at the national level, and weighting factors have been calculated, added to the data file, and applied so that results are representative at the national level.
- Overall, the survey was successfully carried out in 956 clusters.

#### Sampling weights

To account for disproportionate representation in the sample all observations were assigned a sample weight, based on sampling probabilities. More details on the sampling strategy can be found in the Afghanistan 2015 DHS report (7).

#### Inclusion criteria

Women aged 15-49 who had a live birth in the 5 years preceding the survey and provided data on the number of antenatal care (ANC) visits for the most recent live birth.

#### Exclusion criteria

Only women who did not provide data on ANC history were excluded from the study.

### 2.4 Variables

#### 2.4.1 Main Exposure

The main exposure of interest was the number of ANC visits defined as a binary variable categorized as follows:

- 4 and more ANC visits = 0 (4 or more visits)
- Less than 4 ANC visits= 1 (0-3 visits)

#### 2.4.2 Main outcome

The main outcome of interest was neonatal mortality, categorized as a binary outcome as follows:

- Alive or died after 28 days of life = 0
- Died during the neonatal period: born alive and died within the first 28 days= 1

#### 2.4.3 Other variables

Priori confounders included the age of the mother, sex of the child (male or female), marital status, where ANC was attended (health facility or home), ANC provider (health practitioner vs not a health practitioner), place of residence (urban/rural), education of mother, education of father, the wealth index of the family (the two lowest and the two highest quantiles were combined achieving 3 categories) and the smoking status of the mother. To avoid data sparsity, variables that had small observations in a category were regrouped, such as the mother’s education (no education vs some education), marital status (married vs separated or widowed or divorced), and father’s education (no education vs some education). Sex and ANC provider were assessed as priori effect modifiers.

### 2.5 Bias

To minimize results being biased towards over-sampled parts of the population, the complex survey design, such as primary sampling units and sampling weights, were accounted for by using the “svyset” package in STATA.

The survey was translated to Dari and Pashto, and a pretest on participants from the staff of the MoPH and CSO was conducted. Additionally, the interviewers had both male and female representation to limit cultural barriers.

### 2.6 Sample size

To detect a 10% increase among women who attend less than 4 ANC visits in neonatal mortality in comparison to the average neonatal mortality in Afghanistan (45 per 1000 births according to WHO), with 5% significance and 95% power, the total sample size required should be no less than 23,397 women. This calculation was conducted using STATA for sample size calculation in studies testing hypotheses. The sample size provided by the DHS data was 29,461, including all ever-married women aged 15-49 who were either part of the household that was interviewed for the 2015 DHS or visitors who spent the night before the survey was conducted in the household. In the final sample, 19,762 women were included due to missing data on birth history and ANC services.

### 2.7 Statistical analysis

#### 2.7.1 Descriptive statistics

The prevalence of neonatal mortality and the proportion of women in each category who attended less than 4 ANC visits and 4 or more ANC visits were calculated as weighted prevalence and proportions, respectively, taking the survey design into account. Baseline characteristics based on the aforementioned variables were calculated as weighted proportions of the study population, including the proportion of neonates who died and those who didn’t or died after 28 days by age of mother, sex of the child, smoking of mother, where ANC was attended and by who, place of residence, education of mother, education of father, wealth index and smoking status of the mother. For each variable, the weighted prevalence of neonatal mortality was calculated along with 95% CI and p-values.

#### 2.7.2 Preliminary univariate analyses

The association between the number of ANC visits and neonatal mortality was first calculated by cross-tabulation and calculation of the chi-square test for association. Cross tabulation and chi-square test for association were further used to investigate the association between other variables and the outcome of interest (neonatal mortality). Following the same approach, the association between other variables and the exposure of interest (number of ANC visits) was investigated, and any variable associated with both the exposure and the outcome of interest was considered a potential confounder.

#### 2.7.3 Logistic regression analysis

The main outcome of interest was a binary variable; therefore, logistic regression modelling was used for the analysis. The main exposure of interest was categorized into a binary variable: less than 4 ANC visits and 4 or more ANC visits. A crude OR was calculated for the association between ANC visits and neonatal mortality using univariate logistic regression. The odds of neonatal mortality were calculated by the number of ANC visit categories and by each of the aforementioned covariates. For each statistical inference, 95% CI and p-value for the Wald test were obtained. Before conducting the multivariable regression analysis, all covariates were assessed for multicollinearity using the Spearman test, and highly correlated variables were excluded from the model to avoid the effects of multicollinearity on the estimates. ANC location was excluded from the model as it was highly correlated with the ANC provider. ANC provider was kept in the model instead as it showed a stronger association with neonatal mortality (OR: 0.5, 95% CI: 0.36-0.66, p-value<0.001).

Multivariate logistic regression analysis was used to estimate the effect of the number of ANC visits on neonatal mortality after controlling for confounders. The sex of the newborn, mother’s age, mother’s education, father’s education, place of residence, ANC provider, and wealth index were added to the baseline model in sequence to assess their effect on the main association. Each variable that changed the OR of neonatal mortality when ANC was visited for less than 4 days in comparison to those who attended 4 times or more was considered a confounder if it was associated with both ANC visits and neonatal mortality and did not fall on the causal pathway between the number of ANC visits and neonatal mortality. As a result, the mother’s education, place of residence, wealth index, ANC provider, and father’s education were considered confounders.

Additionally, any variable showing variation in the stratum-specific OR was also considered for effect modification. Interaction terms for mother’s education, residence, father’s education, wealth, sex, and ANC provider were added to the model. The adjusted Wald test for joint assessment of interaction terms yielded a low p-value for sex, suggesting significant evidence of interaction. Stratum-specific ORs were derived by adding interaction terms for sex to the main model.

The final model included the number of ANC visits, mother’s education, and place of residence, while sex was tested for interaction. This allowed for the calculation of ORs for neonatal mortality adjusted for all those variables, with 95% CIs and p-values for the Wald test. The p-value for the final model goodness of fit was 0.049, suggesting that this model was the best fit for the data used.

### 2.8 Ethical considerations

The project was approved by the London School of Hygiene and Tropical Medicine ethics committee (reference 30244). Permission to use the dataset was granted directly by the DHS. As per the DHS protocol, all women provided informed consent before answering the survey questions. Most importantly, the dataset did not include any identifiable information about individual participants.

## 3. Results

### 3.1 Participants

In total 25,741 households were selected by the DHS to be included in the survey: 24,941 were occupied, and 24,395 of those occupied were interviewed, representing an overall response rate of 98%. Response rates were lower in urban areas compared to rural areas. There were 30,434 women between the ages of 15 and 45 in the identified households. Interviews were completed with 29,461 of these women, yielding a response rate of 97%. Of the interviewed women, 19,762 provided data on their ANC history for the most recent birth during the five years preceding the survey, out of which 12,141 had ANC for their most recent birth. 9,699 women were excluded from the study due to either not giving birth during the preceding 5 years of the survey or the unavailability of ANC data for the latest pregnancy.

### 3.2 Descriptive data

#### 3.2.1 Baseline characteristics

The characteristics of the study population are presented in Table 1. All proportions presented are weighted to allow for the sampling design of the study. 46.2% of the mothers were 25-34 years old. Infants were fairly distributed by sex and household wealth index. However, due to a few observations in the 4 categorized groups expressing wealth index, the wealth index was regrouped into poor, middle, and rich categories to limit the effect of data sparsity on the estimates. The majority of the mothers had no education, 83%, whereas almost half of the fathers had at least primary or secondary education. Almost all mothers didn’t smoke and were married. The vast majority of the interviewed mothers lived in rural areas (77%). 80% of the women in the sample attended ANC less than 4 times during their pregnancy, and 43.3% of the women had no ANC visits at all. More than half of the mothers received ANC consultations from a health practitioner (59%), and more than half attended a health facility for ANC (58%).

**Table 1.**
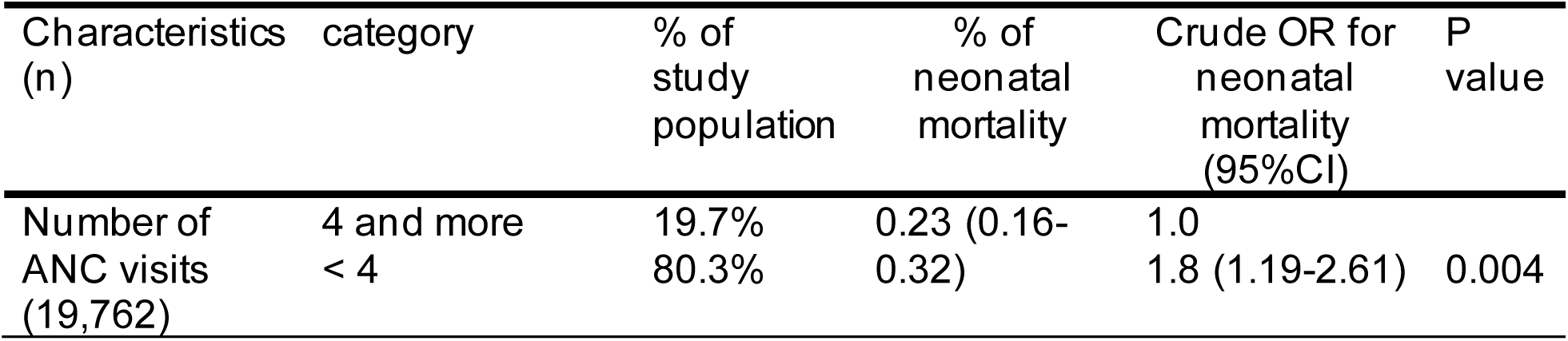
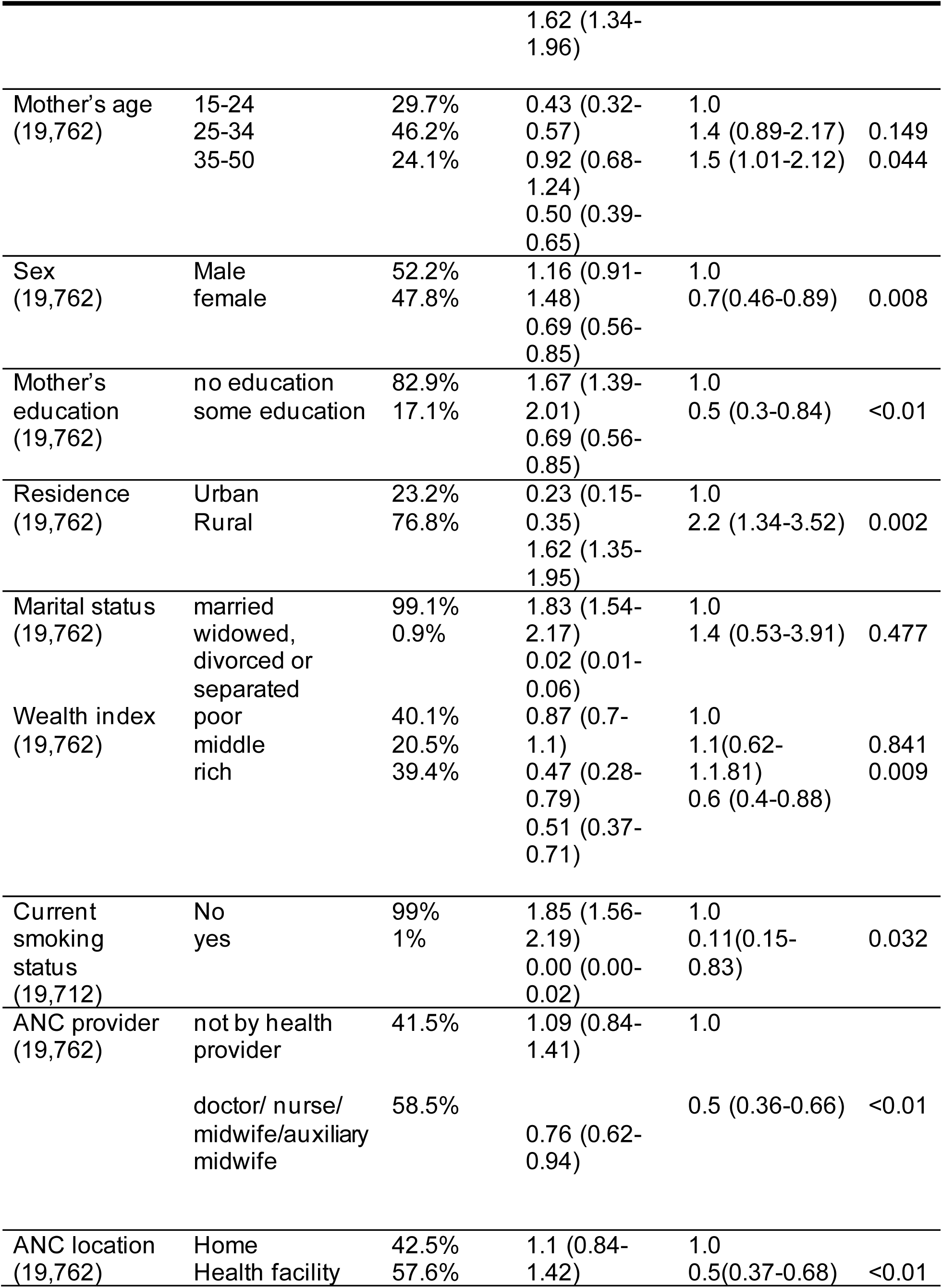
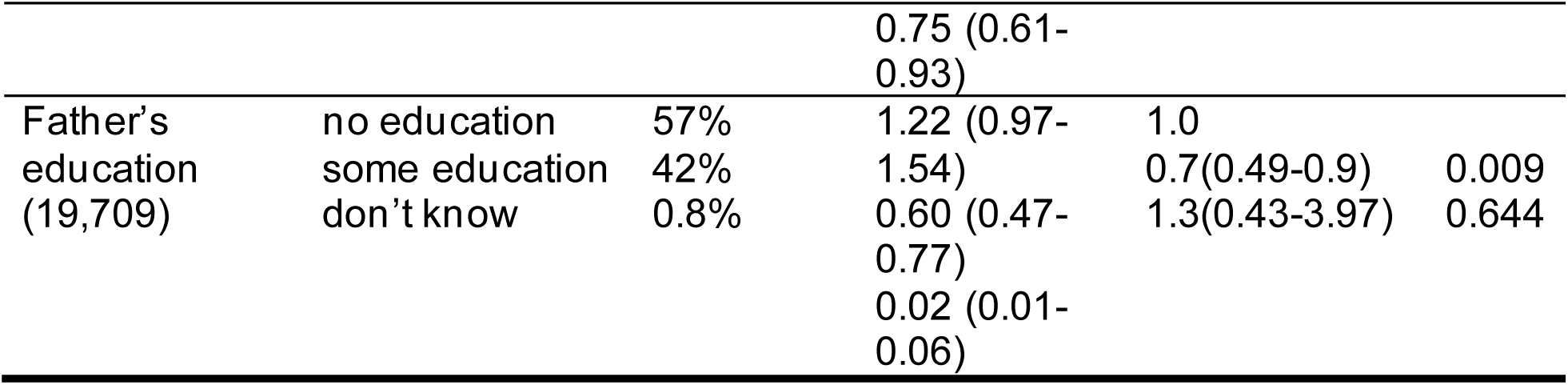
General characteristics of the study population, proportions of neonatal mortality; unadjusted odds ratios with 95% CIs for neonatal mortality; p-values for the association of each factor with neonatal mortality.

#### 3.2.2 Missing data

The response rate on the DHS questionnaire was 98%. Among the 19,762 women who were included in the study, 0.3% of women did not report data on the father’s education, and 0.8% did not know the level of the father’s education. Additionally, 0.3% of women did not report their smoking status. 18 women out of the ones missing data on smoking status did not report the father’s education, while an additional 3 missed data only on their smoking status.

### 3.3 Outcome data

#### 3.3.1 Prevalence of neonatal mortality

The overall prevalence of neonatal mortality in the study population was 1.85% (CI:1.56-2.20, p-value: 0.004). Neonatal mortality was 1.8 times higher in women who did fewer than 4 ANC visits. Women who attended ANC 4 or more times constituted only 20% of the total sample. Neonatal mortality was higher in males (p-value for chi-square association: 0.007), education of the mother (p-value: 0.008), place of residence (p-value: 0.001), ANC provider (p-value: <0.001), ANC location (<0.001), father’s education (p-value: 0.014), and smoking (p-value: 0.0096) however, there were very few observations for women who smoked (1% of the sample), OR: 8.9 in women who smoked (CI:1.2-65.1). Neonatal mortality showed a linear association with wealth index; neonatal mortality was 1.7 times less in richer groups compared to the poor (p-value: 0.009). There was no association shown between neonatal mortality and the current age of the mother; however, in older age groups 35-50 years, some association was expressed in comparison with the youngest age group (OR: 1.5, CI: 1.01-2.12, p-value: 0.04).

#### 3.3.2 Risk factors of neonatal mortality

By cross-tabulation and using logistic regression to assess the association with neonatal mortality, several risk factors for neonatal mortality were identified, as mentioned in Table 1. Mother’s age between 35-50 (OR: 1.5, p-value: 0.04), male sex (OR:1.4, CI: 1.12-2.15, p-value: 0.008), mother not having any previous education (OR:2, CI:1.19-3.38, p-value< 0.001), rural residence (OR:2.2, p-value: 0.002), the ANC provider not being health personnel (OR:2, p-value <0.001), ANC conducted at home (OR: 2, p-value <0.001), father having no previous education (OR: 1.4 in comparison to those who had some education, p-value: 0.009).

#### 3.3.3 Determinants of ANC visits during pregnancy

Table 2 shows the distribution of the study population in each category of binary exposure (less than 4 ANC visits and 4 or more ANC visits). Only 19% of women across all age groups attended ANC 4 or more times. Attendance of ANC was highest in the age group 25-24, with 37% of them attending less than 4 times. ANC attendance was almost fairly distributed by the sex of the child, and 82% of all women in the sample who attended ANC less than 4 times had no education. 77% of women were from rural areas, out of which only 12% attended ANC 4 or more times. ANC was strongly associated with the mother’s education, residence, wealth index, ANC provider, location, and father’s education.

**Table 2.** General characteristics of the study population by number of ANC visits. P-values for the association of each covariate with the number of ANC visits.

| Characteristics<br>(n) | category | % of the study<br>population |  | P-value of<br>association (X<sup>2</sup><br>test) |
| --- | --- | --- | --- | --- |
|  |  | <4 ANC<br>visits | 4 or more<br>ANC visits |  |
| Mother's age<br>(19,762) | 15-24 | 23.7% | 6% | 0.624 |
|  | 25-34 | 37.4% | 8.9% |  |
|  | 35-50 | 19.32 | 4.8 % |  |
| Sex<br>(19,762) | Male | 42 % | 10.2% | 0.723 |
|  | female | 38% | 9.5% |  |
| Mother's<br>education<br>(19,762) | no education | 69 % | 11.2% | <0.001 |
|  | some education | 11.2% | 5.9% |  |
| Residence<br>(19,762) | Urban | 15.4% | 7.8% | <0.001 |
|  | Rural | 64.9% | 11.9% |  |
| Marital status<br>(19,762) | married | 79.6% | 19.6% | 0.0539 |
|  | widowed,<br>divorced or<br>separated | 0.8% | 0.03% |  |
| Wealth index<br>(19,762) | Poor | 35.1% | 5% | <0.001 |
|  | Middle | 17.2% | 3% |  |
|  | rich | 28% | 11% |  |
| Current<br>smoking<br>status<br>(19,712) | No | 80% | 19% | 0.5039 |
|  | yes | 0.8% | 0.2% |  |
| ANC provider<br>(19,762) | not by health<br>provider | 40.6% | 0.8% | <0.001 |
|  | doctor/ nurse/<br>midwife/auxiliary<br>midwife | 39.7% | 18.8% |  |
| ANC location<br>(19,762) | Home | 41.3% | 1.1% | <0.001 |
|  | Health facility | 39% | 18.5% |  |
| Father's education (19,709) | no education | 48% | 9.1% |  |
|  | some education | 31.6% | 10.5% |  |
|  | don't know | 0.7% | 0.1% | <0.001 |

### 3.4 Main results

#### 3.4.1 Univariate analysis

As shown in Table 3, the crude OR for the association between the number of ANC visits and neonatal mortality was 1.8 (CI:1.19 −2.61, p-value:0.004) comparing women who attended ANC less than 4 times to those who attended ANC 4 or more times.

**Table 3.**
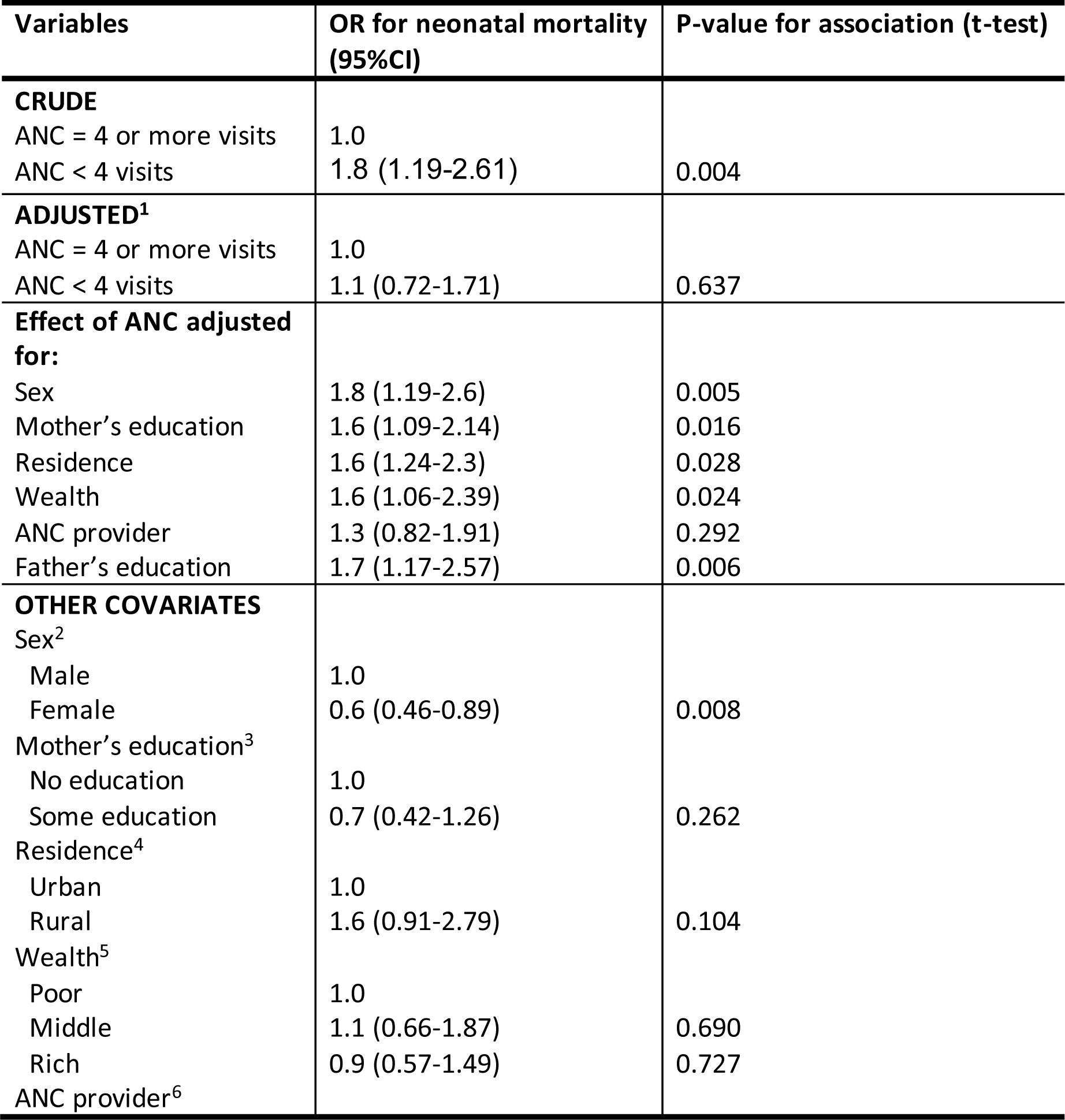
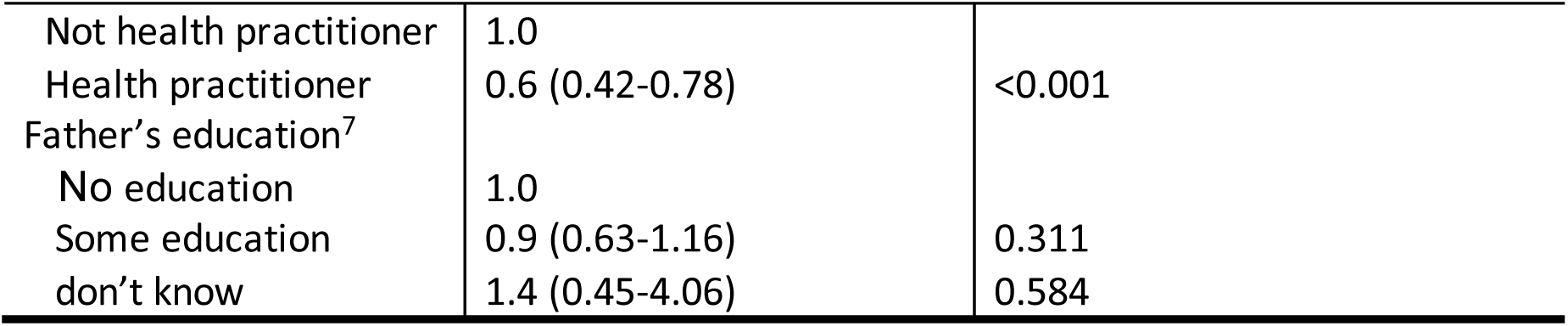
Logistic regression: crude and adjusted odds ratios for the association between the number of ANC visits and neonatal mortality.

#### 3.4.2 Multivariable logistic regression analysis

Covariates that showed a strong association with the number of ANC visits and neonatal mortality were included in the multivariable logistic regression analysis. After adjusting for sex, mother’s education, residence, ANC provider, and father’s education using multivariable logistic regression (table 3), the odds of neonatal mortality in those whose mothers attended ANC less than 4 times (adjusted OR 1.1; 95% CI 0.72-1.71; p-value=0.628), suggested overall confounding however, there was no enough evidence against the null hypothesis of the OR being 1 comparing women who attended less than 4 times to those who attended 4 times or more (high p-value and the CI including the null of 1). Despite not showing any significant association between ANC visits and neonatal mortality after adjusting for potential confounders, the OR decreased by 39%. After adjusting for each covariate at a time and comparing the adjusted OR with the crude OR for the association between neonatal mortality and the number of ANC visits, mother’s education decreased the OR by almost 11% from 1.8 to 1.6 (95% CI: 1.1-2.42, p-value: 0.016), place of residence decreased the OR by 11% from 1.8 to 1.6 (95% CI: 1.1-2.32, p-value:0.028), wealth index decreased the OR by 11% from 1.8 to 1.6 (95% CI: 1.06-2.39, p-value: 0.024), and father’s education decreased the OR by 6% from 1.8 to 1.7 (95% CI: 1.2-2.57, p-value: 0.006). The ANC provider decreased the OR by 28% to 1.3, but did not yield enough evidence against the null hypothesis of the OR being 1, and the CI included the null of 1. As a result, the mother’s education, place of residence, wealth index, ANC provider, and father’s education were considered confounders of the main association since they are associated with both the number of ANC visits and neonatal mortality and do not fall on the causal pathway between the two, and their inclusion in the model affected the crude OR.

Multivariable logistic regression also yielded estimates for the other variables that were found to be associated with neonatal mortality. After adjusting for all other variables in the study, including the exposure variable (number of ANC visits), the odds of neonatal mortality were 15% higher in females (p-value:0.008), 23% higher in rural areas (p-value:0.028), and 24% lower when the ANC provider was a health practitioner (p-value: <0.001).

#### 3.4.3 Effect modification

The final logistic model for the association between ANC and neonatal mortality included the mother’s education and place of residence as a best fit for the data provided. Using a model including all confounders for the relationship, covariates were tested one at a time for possible effect modification using an interaction term. By adding the interaction term for sex into the model, the adjusted Wald test for joint interaction assessment yielded a p-value of 0.032, showing possible effect modification by sex.

#### 3.4.4 Stratum-specific effects

An interaction term was added to the main model using the identified effect modifier at a time to derive stratum-specific ORs. When ANC was attended 4 or more times, the odds of neonatal mortality were 1.4 times in females compared to males. In male newborns, the odds of neonatal mortality were 2.6 times higher when ANC was attended less than 4 times compared to when it was attended 4 or more times.

**Table 4.**
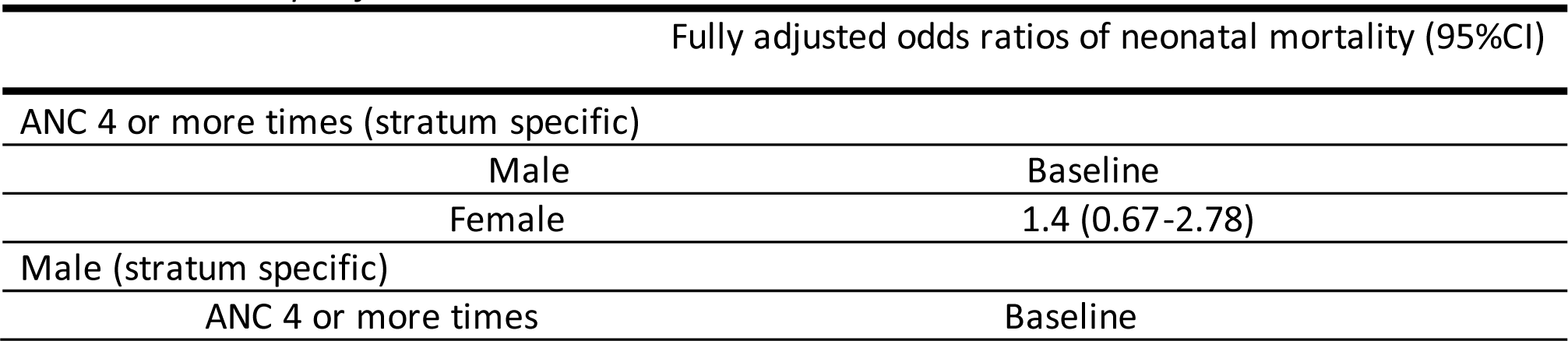
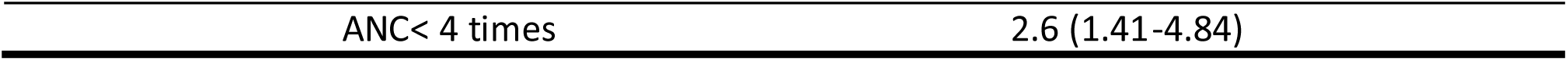
Stratum-specific ORs.

## 4. Discussion

### 4.1 Principal findings

The overall prevalence of neonatal mortality in the used sample was 2% of the entire sample size. The prevalence of neonatal mortality was 86% higher in women who attended ANC less than 4 times (1.62, CI:1.34-1.96) in comparison to those who attended ANC 4 or more times in Afghanistan. This study revealed that the number of ANC visits is strongly associated with neonatal mortality. The odds of neonatal mortality were 1.8 times higher when the mother went for fewer than 4 ANC visits during her pregnancy. However, after adjusting for the sex of the child, mother’s education, place of residence, ANC provider, wealth and father’s education, this association almost disappeared as the odds dropped to 1.1, but the confidence interval included 1, and the p-value for the null hypothesis of no association between neonatal mortality and the number of ANC visits after adjusting for the mentioned covariates was 0.628. In the multivariable regression model, this study identified several risk factors for neonatal mortality. Male babies were 1.6 times more likely to die in the neonatal period, women living in rural areas were 1.7 times more likely to face neonatal mortality, and it was 1.8 times more likely for the child to die in the neonatal period if ANC was conducted by someone who was not a health practitioner. The wealth index only showed an association with neonatal mortality in rich groups and, therefore, was excluded from the final model to avoid inaccurate estimates. This study also revealed the significant role of the mother’s education, place of residence, father’s education, and ANC provider as confounding factors in the main relationship and the role of the newborn’s sex as an effect modifier for the main relationship. The interactive effect of sex yielded that when attending ANC 4 or more times, it was 1.4 times more likely for the newborn to die during the first 28 days of life if the sex of the newborn was female. This effect was reversed when ANC was attended less than 4 times as the newborn was 2.6 times more likely to die during the first 28 days if the sex was male, showing that in situations of ANC attendance less than the minimum recommended times, female sex played a protective role against neonatal mortality and males became more likely to die than females in the neonatal period.

### 4.2 Strengths and limitations

#### 4.2.1 Strengths

There are several strong points in this study. The study was based on the DHS data, which uses a standardized methodology to facilitate cross-sectional analysis of data from a nationally representative sample of the population of Afghanistan. The DHS uses internationally reliable definitions; therefore, the collected data on ANC, wealth index, and other variables used in this study can be trusted, and the results can be compared with other studies on the same topic. The complex survey design of sampling and weighting used in the DHS was taken into account in the statistical analysis, allowing for disproportionate representation in the sample and ensuring more accurate estimates and confidence intervals. Moreover, some known risk factors for neonatal mortality, such as sex, and potential confounders for the association between the number of ANC visits and neonatal mortality, such as the education level of the parents, were considered in the analysis that revealed a more in-depth and accurate understanding of their effect as confounders and effect modifiers.

#### 4.2.2 Limitations

A major limitation of this study is treating ANC visits of any more than 4 and any less than 4 equally, possibly underestimating the real protective effect of the increased number of ANC visits and its dose-response relationship with neonatal mortality. Similarly, conducting no ANC visits (43.3% of the sample) was treated as conducting 1-3 visits; therefore, women who did not conduct ANC visits were overrepresented in the sample and this category.

##### Study design

The causal effect between the number of ANC visits and neonatal mortality cannot be ensured with a cross-sectional study design since the data regarding the exposure and the outcome of interest were collected at the same point in time.

##### Sample size

Despite this study including 29,461 interviewed women, the final sample used for the analysis only included 19,762 women who provided data on ANC history of the last birth that occurred during the 5 years preceding the survey, therefore reducing the sample size by 17% from the minimum sample size that was estimated for a 5% significance and 95% power (23,397). Nonetheless, this estimation was based on assuming a 10% increase in the prevalence of neonatal mortality from the country average, which did not occur when analyzing the prevalence of neonatal mortality in the used sample. In the used sample, the overall prevalence of neonatal mortality was 2%, and the prevalence in women attending ANC less than 4 times was 19% higher than those who attended 4 or more times.

##### Random error

The analysis followed a 5% significance, suggesting that there is a low chance of false positive results. However, considering the sample size and the fact that the variables were carefully selected for analysis and did not exceed 10 in total, unnecessary analysis was avoided, and as a result, the likelihood of random error decreased.

##### Bias

Selection bias was limited in this study by using the two-stage sampling frame, which allowed for a representative sample to be interviewed, the household listing operation, the replacement clusters for inaccessible areas, and with a 97% response rate on the women’s questionnaire, and most respondents completing the questionnaire without refusing questions. However, response rates were lower in urban areas compared to rural areas. This mostly accounted for the response rates of the men’s questionnaire, as they were mostly absent from the household at the time of the data collection. Additionally, no replacement of pre-selected households was allowed in the implementation period. To further limit selection bias, the DHS interviewers entered some inaccessible areas from neighboring countries where context-specific challenges could affect access to ANC.

The data collected on both ANC and neonatal mortality are subject to misclassification bias as they were self-reported, and reporting errors might have occurred due to the interviewees’ misunderstanding of the questions. A woman might have visited the doctor or a community worker for a health issue related to something other than antenatal care and counted it as an ANC visit, and neonatal mortality, similarly, could be misclassified in terms of the age of the newborn at death or mistaken with stillbirth. Stillbirth and neonatal mortality are considered widely underreported due to either misclassification or recall bias, as mothers are less likely to recall children who died very early in their lives. Recall bias is a major limitation in this study design, which the DHS mitigated by collecting data on the most recent birth over the preceding 5 years, not to mention that data on other covariates, such as father’s education, ANC provider, and location, mostly relied on past information. The age of the mother was reported as the date of birth and later calculated to avoid recall bias; however, it could not be linked with the age of the mother at the time of the pregnancy, yielding a less accurate implication of the mother’s age on ANC and consequently the outcome of the pregnancy. Information bias was minimal considering the quality insurance measures that were applied during the process of data collection by the DHS team, such as training the field staff on the questionnaire and having a separate team do the data entry to avoid confirmation bias while compiling the data. Finally, due to the overrepresentation of women who did not have any ANC visits, the final estimates might be biased towards that group.

##### Missing data

Some data on the father’s education and smoking were missing. These few missing values could not have significantly affected the accuracy of the estimates. Non -smoking was highly overrepresented in the sample (99%), therefore biasing the results of the effect of smoking on the main relationship. However, smoking was not included in the final model.

##### Residual confounding

There was not much missing data; therefore, there is a very small possibility that residual confounding from variables with missing data might have affected the results. However, many other potential confounders for the relationship that are known risk factors of neonatal mortality and could have also been associated with the number of ANC visits were not included in this study, such as age at birth, place of delivery, cesarean section, and unsafe security situation, for which data were not available. These unassessed factors could have caused residual confounding.

### 4.3 Strengths and weaknesses in relation to existing literature

Despite many studies on determinants of neonatal mortality globally, in low and middle-income countries (17,18,19,20) and specifically in Afghanistan (11), few studies have investigated the relationship between ANC and neonatal mortality in Afghanistan. This study explored a new aspect of the importance of a minimum of 4 visits as a true game changer. Within the complex crisis-related context of Afghanistan, this study is the first to assess the added value of promoting more than 4 ANC visits to prevent neonatal mortality. In a study analyzing the national health survey of 2018 in Afghanistan, the contents of ANC services were assessed (14). The study found that ANC was underutilized, and those who had more knowledge about pregnancy complications were more likely to utilize these services. Ultimately, it was concluded that not only the type and components of ANC services but also increasing knowledge would improve attendance. Our study had an additional value that complements this study, highlighting the importance of education and its influence on the number of ANC visits and neonatal mortality. In a study investigating the impact of inadequate ANC on neonatal survival in Rwanda, (21) it was concluded that uptake of ANC improved newborn survival and addressing the quality of ANC by skilling health workers can potentially contribute to improving neonatal outcomes, in parallel our study showed that having a skilled health worker providing ANC was associated with both the number of visits and neonatal mortality pointing at a possible confounding effect and therefore assuring that scaling up access to skilled health workforce will not only improve the adherence of women to conduct more than 4 visits but also decrease neonatal mortality. An extensive study on the determinants of neonatal mortality in Afghanistan identified several individual, maternal, and community-level factors that influence neonatal mortality, including utilization of ANC services (11). This study complements the findings while emphasizing the importance of encouraging women to adhere to at least 4 visits to decrease th e odds of neonatal mortality by almost 86%.

### 4.4 Interpretation of the study

This cross-sectional study revealed that there is an almost doubled risk of neonatal mortality when ANC is attended less than 4 times in comparison to when it is attended 4 or more times, which could be interpreted as increased importance of pregnancy monitoring as the pregnancy progresses and towards the later stages to detect possible high-risk deliveries caused by conditions such as placental disorders or disruptions or intraamniotic infections and failed treatment of urinary tract infections. This aspect does not rule out the importance of pregnancy monitoring in the early stages, which is crucial for neurological development and avoiding preventable poor pregnancy outcomes. It is well known that premature birth, birth complications (asphyxia and trauma), and neonatal infections remain the leading causes of neonatal deaths, followed by congenital anomalies, and adequate pregnancy follow-up towards the third trimester remains key to early detection of such conditions and preventing negative outcomes. Another possible interpretation is that committing to regular and more frequent ANC visits improves with an increased number of ANC visits. The more the pregnant woman visits, the more aware she is of alarming symptoms that require the consultation of health personnel, and it is more likely to be timely diagnosed and treated for morbidities that could affect the pregnancy outcome. However, given that this study focused on the cutoff point of 4 visits, it highlights the significant added value of maintaining no less than 4 ANC visits, which is aligned with the WHO recommendations and should be consistently encouraged via all health policies, even in conflict-affected settings such as Afghanistan. Future studies could assess for the possible confounding and effect modification by security situation, as women in areas that were undergoing deteriorated security unrest could have been less likely to move freely and safely to attend ANC services and therefore possibly attended less and had worse pregnancy outcomes.

### 4.5 Generalizability

The findings from this study are generalizable to the population of Afghanistan due to having a representative sample covering different provinces, even those that were hard to reach and were accessed from neighboring countries; however, the security situation did affect access to some areas that were considered unsafe. These findings could be extrapolated to other LMICs where resources are unequally distributed and also in conflict-affected settings. Nevertheless, we should be cautious in our interpretation as it might not be fully applicable to conflict-affected settings where disrupted security situations can potentially affect access to health care. However, a general assumption can be made that worse outcomes can be expected in such settings.

### 4.6 Unanswered questions and future research

This study highlighted the importance of maintaining at least 4 ANC visits, which has a significant impact on preventing neonatal mortality. It addressed some major confounding and interactive factors that play a key role in manipulating this relationship, such as the education of parents and the sex of the newborn. However, this study does not explain the dose-response relationship that the number of ANC visits might have with neonatal mortality. A study investigating whether there is a linear relationship between the two and controlling for other potential confounders, such as security situation, distance from the health facility, weight of the newborn, preterm birth, age of the mother during pregnancy, around birth, and postnatal factors, is suggested. To better understand the causal relationship, a longitudinal study design could be used in future studies in which pregnant women are followed up during their pregnancy and the outcomes of the pregnancy are witnessed in real-time, limiting recall bias. Furthermore, the confounding effect of wealth should be further investigated using a bigger sample in which women whose neonates died are more represented, in addition to the effect of ANC being provided by a health practitioner, which could have interactive effects on the main relationship.

## 5. Conclusion and recommendations

This study showed that neonatal mortality was 1.8 times higher when women attended ANC less than 4 times in comparison to those who attended 4 times or more. While the study’s cross-sectional design limits assumptions about causality, it explained the importance of attending at least 4 ANC visits, ultimately aligning with the WHO recommendations. In conclusion, it is suggested that raising awareness on adhering to a minimum of 4 visits would have a significant impact on neonatal survival and cannot be compromised in LMICs and conflict-affected settings such as Afghanistan. The findings of this study highlight the need to improve ANC attendance and strengthen access to antenatal care in conflict-affected settings such as Afghanistan. To improve these aspects, health education sessions, community-based activities, and the introduction of cross-cutting health policies are crucial. The existing governmental bodies and humanitarian organizations working in such conflict settings should prioritize the availability of ANC services and access for communities in hard-to-reach and rural areas with mobile health clinics. On the other hand, efforts should be made to consolidate national healthcare policies, focusing on strategies to improve ANC coverage and address disparities between rural and urban areas. In the long term, advocacy for international funding to rebuild the disrupted health system should be considered to build the existing healthcare system’s resilience. Community engagement activities remain key to delivering knowledge and raising awareness aimed at improving the perception of women and their family members regarding antenatal care and its role in neonatal survival. Moreover, these activities could contribute to building the trust of the communities in the health system by ensuring their active participation in designing and delivering effective interventions to meet their needs.

## Data Availability

The data is available through the Demographic and Health Surveys platform upon request for academic and research purpose.

https://dhsprogram.com/Data/

## Footnotes

1 Adjusted for sex, mother’s education, residence, wealth, ANC provider, and father’s education

2 Adjusted for ANC, mother’s education, residence, wealth, ANC provider, and father’s education

3 Adjusted for ANC, sex, residence, wealth, ANC provider, and father’s education

4 Adjusted for ANC, sex, mother’s education, wealth, ANC provider and father’s education

5 Adjusted for ANC, sex, residence, mother’s education, ANC provider and father’s education

6 Adjusted for ANC, sex, mother’s education, residence, wealth and father’s education

7 Adjusted for ANC, sex, mother’s education, residence, and ANC provider

